# Natural Language Processing for the Ascertainment and Phenotyping of Left Ventricular Hypertrophy and Hypertrophic Cardiomyopathy on Echocardiogram Reports

**DOI:** 10.1101/2023.05.17.23290116

**Authors:** Adam N. Berman, Curtis Ginder, Zachary A. Sporn, Varsha Tanguturi, Michael K. Hidrue, Linnea R. Borden, Yunong Zhao, Ron Blankstein, Alexander Turchin, Jason H. Wasfy

**Affiliations:** Division of Cardiovascular Medicine, Department of Medicine, Brigham and Women’s Hospital, Harvard Medical School, Boston, MA; Department of Medicine, Massachusetts General Hospital, Harvard Medical School, Boston, MA; Cardiology Division, Department of Medicine, Massachusetts General Hospital, Harvard Medical School, Boston, MA; Division of Performance Analysis and Improvement, Massachusetts General Physicians Organization, Massachusetts General Hospital, Harvard Medical School, Boston, MA; Division of Endocrinology, Department of Medicine, Brigham and Women’s Hospital, Harvard Medical School, Boston, MA

**Author notes:** <u>Address for Correspondence:</u> Jason H. Wasfy, MD Mphil, Massachusetts General Hospital, 55 Fruit Street, Boston, MA 02115.

**Keywords:** Left ventricular hypertrophy, natural language processing, hypertrophic cardiomyopathy, LVH-associated diseases

## Abstract

**Objective:** Extracting and accurately phenotyping electronic health documentation is critical for medical research and clinical care. While there are a variety of techniques to accomplish this task, natural language processing (NLP) has been developed for numerous domains to transform clinical documentation into data available for computational work. Accordingly, we sought to develop a highly accurate and open-source NLP module to ascertain and phenotype left ventricular hypertrophy (LVH) and hypertrophic cardiomyopathy (HCM) diagnoses on echocardiogram reports from a diverse hospital network.

**Methods:** 700 echocardiogram reports from six hospitals were randomly selected from data repositories within the Mass General Brigham healthcare system and manually adjudicated by physicians for 10 subtypes of LVH and diagnoses of HCM. Using an open-source NLP system, the module was developed on 300 training set reports and validated on 400 reports. The sensitivity, specificity, positive predictive value, and negative predictive value were calculated to assess the discriminative accuracy of the NLP module.

**Results:** The NLP demonstrated robust performance across the 10 LVH subtypes with overall sensitivity and specificity exceeding 96%. Additionally, the NLP module demonstrated excellent performance detecting HCM diagnoses, with sensitivity and specificity exceeding 93%.

**Conclusion:** We designed a highly accurate NLP module to determine the presence of LVH and HCM on echocardiogram reports. Our work demonstrates the feasibility of NLP to detect diagnoses on imaging reports, even when described in free-text. These modules have been placed in the public domain to advance research, trial recruitment, and population health management for individuals with LVH-associated conditions.

## 1) Introduction

Extracting and accurately phenotyping documentation from electronic health records (EHRs) is incredibly valuable. In medical research, this work can identify subjects eligible for clinical trials, build observational cohorts to study clinical outcomes and/or disparities, and assess for the population burden of specific conditions. In clinical care, this work can facilitate quality assurance or create clinical registries for population health management. While there are a variety of techniques used to extract and phenotype EHR documentation, natural language processing (NLP) has been used in numerous clinical applications^1-11^ and continues to be developed across various critical domains to transform clinical documentation into data available for research and clinical care.^10,11^

One particular area that is apt for NLP development is phenotyping clinical imaging reports across different hospitals and reading physicians. Imaging reports often contain a vast amount of actionable and clinically relevant information that can be harnessed for advancing patient care and research. Although recent advances in the EHR have enabled the categorization and storage of certain imaging report templates automatically within structured data fields, many aspects of reporting remain free-text and fully customized by the interpreting clinician. In fact, there are many types of imaging reports that require the flexibility of free-text descriptions to effectively communicate important clinical details. Additionally, access to structured data may be limited or unavailable for regular research or operational purposes. Accordingly, there is a significant opportunity to build accurate NLP tools to accurately and efficiently phenotype clinical imaging reports.

Echocardiogram reports contain detailed descriptions about myocardial structure and function. Reporting templates with structured data are gathered inconsistently in different echocardiography laboratories and echocardiographers often add free-text descriptors to better detail their clinical observations. Additionally, structured data fields may be changed over time or may not have been in use for older reports. One clinically important echocardiographic finding that may be described in free-text form is that of left ventricular hypertrophy (LVH). LVH is a common finding and its accurate classification can lead to important clinical decisions by the referring clinician. Similarly, echocardiographic findings consistent with hypertrophic cardiomyopathy (HCM) may also be described in free-text to inform downstream clinical decision making.

Accordingly, we set out to design and validate an open-source NLP module to ascertain and phenotype LVH and diagnoses of HCM from the text of echocardiogram reports. Our aim was to develop a highly accurate module built from multiple echocardiogram databases within the Mass General Brigham (MGB) hospital system that would ultimately enable the large-scale analysis of subjects with echocardiographic LVH and HCM. By developing an open-source module, our aim was to broadly advance research and clinical operations related to LVH-associated diseases.

## 2) Materials and Methods

We sought to develop an NLP module to detect the presence and subtype of LVH and the finding of HCM on echocardiogram reports. Based on our clinical expertise and the input of an advanced echocardiographer, we designed the module to detect and classify 10 subtypes of LVH, as detailed in **Figure 1**. Additionally, we further designed the module to detect whether the echocardiogram report referenced a likely diagnosis of HCM. The module was designed to assess for diagnostic language for these LVH subtypes and HCM on a phrase-by-phrase and sentence-by-sentence level. For example, a report stating that, “There is evidence of mild concentric increased LV wall thickness” would be diagnostic of output #3 (mild concentric LVH). In contrast, a report stating that, “Moderate asymmetric left ventricular hypertrophy is present” would be diagnostic of output #7 (moderate asymmetric LVH). The NLP module was designed to flexibly and accurately recognize varying sentence structures and categorize reports according to their LVH subtype and HCM. Ultimately, our aim was to develop NLP algorithms that recognize the diverse linguistic formulations that echocardiographers employ in clinical reports to describe the findings of LVH and HCM.

**Figure 1:**
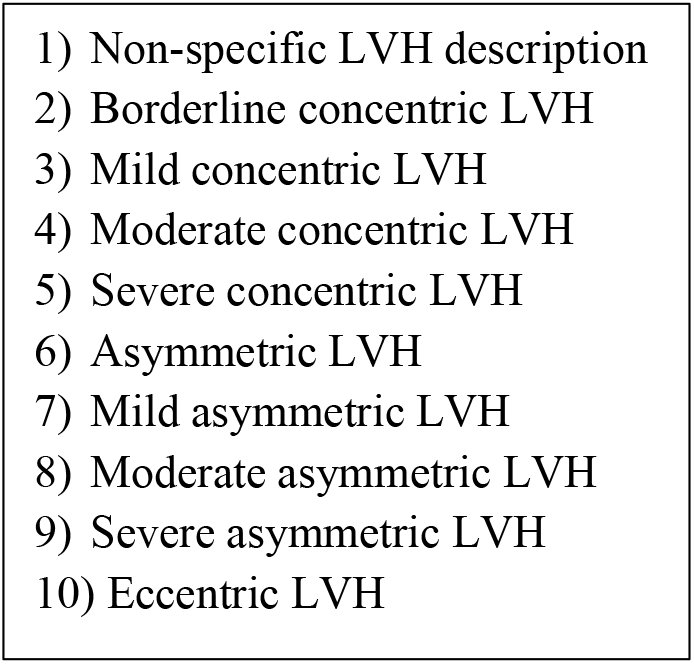
Natural Language Processing Derived LVH Subtypes:

### 2.1) Document Selection

Full text-based echocardiogram reports were extracted from the MGB Research Patient Data Registry (RPDR)^12^ which represents a centralized data warehouse that consolidates clinical data from sites of care within the MGB network. MGB is a large, multi-institutional and integrated healthcare delivery network serving the greater Boston, MA area and includes 5 academic and 6 community hospitals along with numerous associated outpatient clinics. MGB has approximately 2.5 million unique clinical encounters per year and has more than 800,000 patients receiving primary care. A query of the RPDR system in December 2022 demonstrated that there were ∼1.1 million text-based echocardiogram reports available from 1/1/2000 through 12/1/2022.

The NLP module was initially developed on an unselected contemporary set of approximately 17,250 unique echocardiogram reports from approximately 15,000 patients within the MGB system occurring from 1/1/2017. After numerous iterative rounds of design and beta testing, we began the formal validation process in October 2022.

For this process, we queried the well-established Massachusetts General Hospital (MGH) echocardiogram database for individuals having transthoracic echocardiograms with interventricular septal dimensions of ≥12 mm and/ or posterior wall dimensions of ≥12 mm. Separately, we directly queried the RPDR system for individuals with ≥ 2 International Classification of Diseases (ICD) codes for diagnoses that were likely to yield echocardiograms with LVH. The queried ICD codes included those for hypertensive heart disease, cardiomegaly, and hypertrophic cardiomyopathy. The goal of this process was to ultimately generate a set of echocardiogram reports with a moderate concept prevalence of LVH and HCM to be able to calculate the module’s performance.

After obtaining the echocardiogram reports from individuals meeting either of these specifications, we then selected a random set of 700 full-text echocardiogram reports from 700 unique individuals. Of these, 300 were randomly selected for use as a training set and the remaining 400 were set aside for the validation set. The 400 validation set echocardiogram reports represented reports from two academic medical centers, four community hospitals, and seven associated outpatient satellite clinics. The designer of the NLP module (ANB) was blinded to the validation set. This study was approved by the Institutional Review Board at MGB and was granted a waiver of informed consent.

### 2.2) Adjudication

All 700 echocardiogram reports were manually adjudicated for the presence / subtype of LVH and language diagnostic for HCM by a trained physician adjudicator (ZAS) who was not involved in the development of the NLP. The physician adjudicator was instructed to label each echocardiogram based on the highest ordinal level of LVH (for subtypes 1-10 as per **Figure 1**). Accordingly, as is typical with echocardiogram reports, there may be multiple references within a given report to the concept of LVH. The first reference may state in a general fashion that, “Left ventricular wall thickness is increased.” Only later, in the conclusion portion of the report, there may be a second reference specifying that the echocardiogram demonstrates, “Moderate asymmetric septal hypertrophy.” In such an echocardiogram report, the adjudicator was instructed to select subtype 8 as this was the highest ordinal LVH category reported in the echocardiogram text.

For the 400 validation set echocardiogram reports, two cardiologists (CG and JHW) reviewed all reports that contained discrepancies between the NLP output and the manual adjudication to determine the finalized gold-standard manual adjudication. See **Table 1** for the number of unique report-level references to each LVH subtype and HCM in the 400 validation set reports. Since our analytic intent was to create NLP to detect LVH subtypes and HCM in echocardiogram text reports, we did not re-adjudicate primary echocardiographic images.

**Table 1:** Number of unique report-level references to each LVH subtype and hypertrophic cardiomyopathy in the 400 validation set echocardiogram reports

| Subtype | Number of report-level references | Concept prevalence |
| --- | --- | --- |
| Non-specific LVH description | 93 | 23.3% |
| Borderline concentric LVH | 9 | 2.3% |
| Mild concentric LVH | 54 | 14.0% |
| Moderate concentric LVH | 15 | 3.8% |
| Severe concentric LVH | 5 | 1.3% |
| Asymmetric LVH | 17 | 4.3% |
| Mild asymmetric LVH | 12 | 3.0% |
| Moderate asymmetric LVH | 6 | 1.5% |
| Severe asymmetric LVH | 5 | 1.3% |
| Eccentric LVH | 0 | 0% |
| Hypertrophic cardiomyopathy | 29 | 7.3% |

### 2.3) NLP Development

The NLP module was created using the open-source Canary NLP platform.^5,7,10,11,13-16^ We elected to use this NLP platform for the following reasons: (1) it implements NLP algorithms transparently, thereby facilitating tailored optimization and error correction; (2) it is easily transferrable to other datasets and institutions; and (3) it has been demonstrated to achieve higher accuracy than other NLP techniques.^16^

For the NLP module, a customized set of *word classes* was created based on prior clinical experience as well as initial beta testing. *Word classes* contain sets of linguistically-related words or phrases that can be used to create more complex sentence structures. A simplified set of *word classes* (denoted by the presence of “>“) is demonstrated in **Figure 2**.

**Figure 2:**
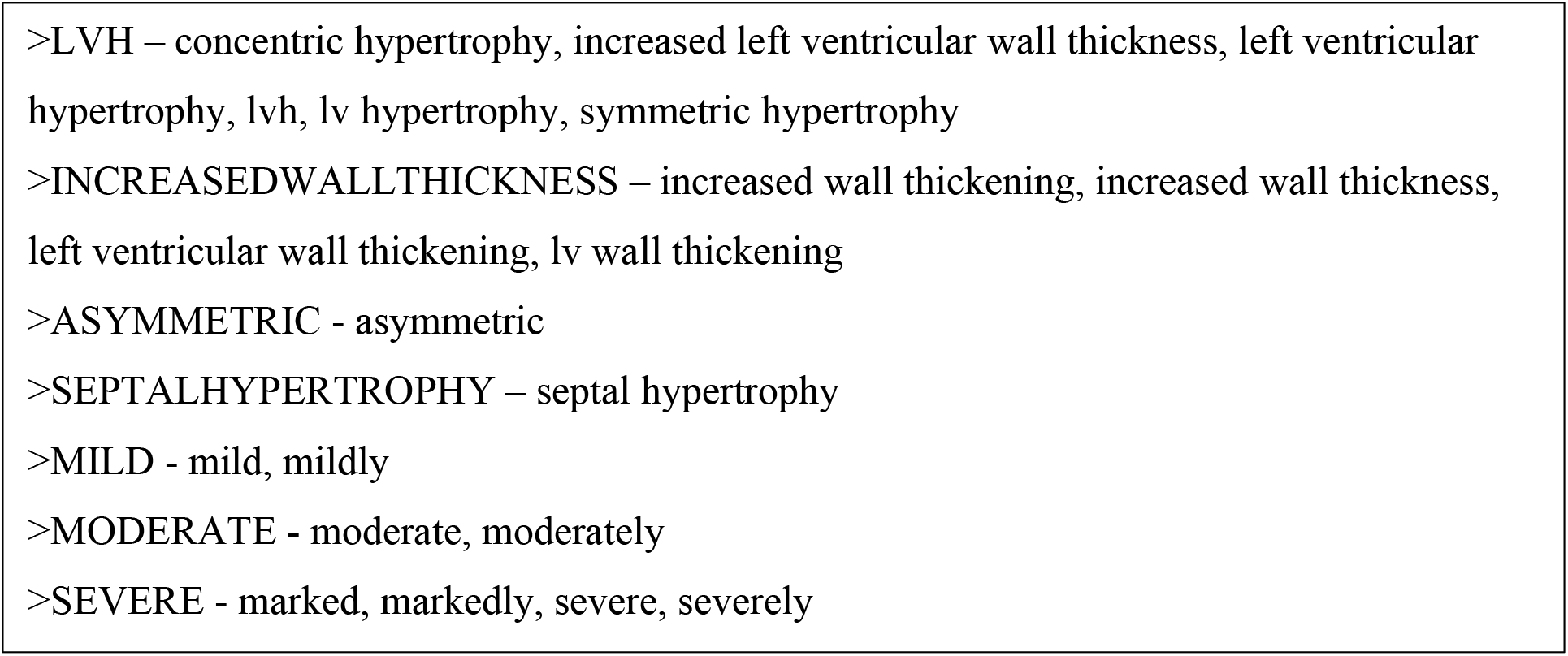
A simplified set of *word classes* which are used to build sentence structures

*Phrase structures* were then customized from *word classes* to create meaningful elements of linguistic information which could then be extracted as numbered outputs for analytic work. An example *phrase structure* to classify a sentence such as, “The findings of severe asymmetric septal hypertrophy are consistent with hypertrophic cardiomyopathy” is shown in **Figure 3**. This example sentence would then resolve to the numbered output 9 for “severe asymmetric LVH” and the output indicating HCM. In this NLP module, there are 27 distinct *word classes* and nearly 500 unique *phrase structures* to accurately and flexibly categorize variations in linguistic formulations.

**Figure 3:**
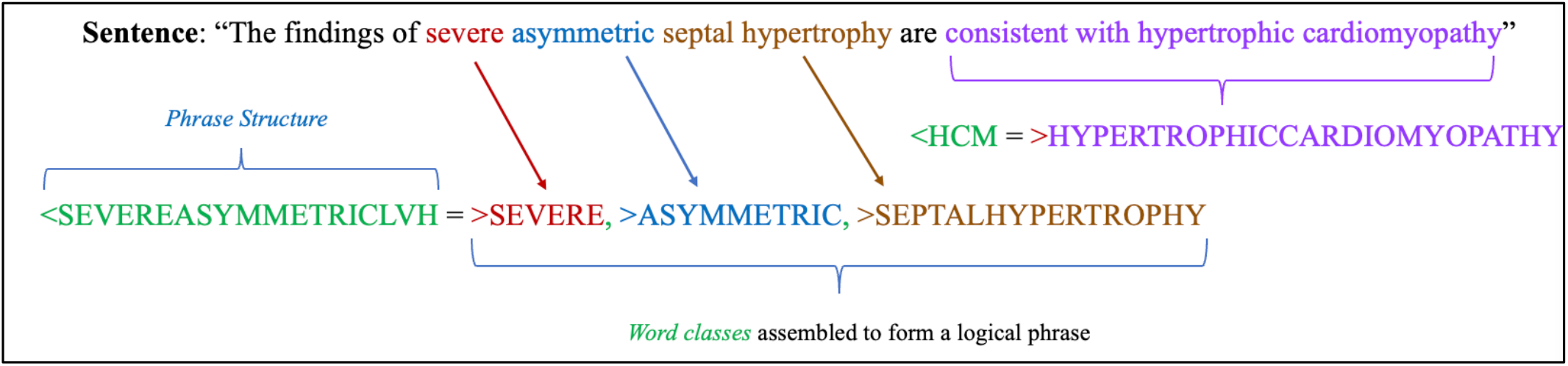
An example of a *phrase structure* to classify a sentence

In addition to categorizing references to LVH and HCM, the NLP module was built to exclude references to negations or intentionally non-diagnostic phrase structures. Accordingly, a sentence stating, “There may be evidence of LVH” or one that states, “No evidence of hypertrophic cardiomyopathy” were programmed to be disregarded by the NLP system.

### 2.4) Performance Calculations

We calculated the sensitivity, specificity, positive predictive value (PPV), negative predictive value (NPV), and the accuracy – collectively, the “performance” – for the NLP module based on each echocardiogram report, referenced to the “gold standard” manual adjudication. Accuracy was defined as (true positives + true negatives) / (true positives + false positives + false negatives + true negatives). We defined an NLP output as a true positive when the NLP output was an <u>exact</u> numeric match with the manual adjudication. Accordingly, if the NLP output for a given echocardiogram indicated the presence of mild concentric LVH (output #3) but the manual adjudication indicated only the presence of non-specific LVH (output #1), this was conservatively categorized as a false negative.

## 3) Results

Out of 400 echocardiogram reports in the validation set, 216 reports had diagnostic references to LVH, generating a concept prevalence of 54%. Additionally, there were 29 reports with a diagnostic reference of HCM, yielding a concept prevalence of 7.3%, **Table 1**.

The NLP module demonstrated robust performance across LVH outputs with sensitivity and specificity exceeding 96%. The PPV for the LVH outputs was 99.5% (95% CI 97.4-99.9%) and the NPV 96.3% (95% CI 92.6-98.5%). Similarly, the NLP module demonstrated excellent performance for the HCM output, with sensitivity and specificity exceeding 93%. The PPV for the HCM output was 93.1% (95% CI 77.2-99.2%) and the NPV was 99.5% (95% CI 98.1-99.9%). The full performance details of the module are demonstrated in **Table 2** and **Table 3**.

**Table 2:** Performance of the Natural Language Processing Module within the LVH Outputs

|  |  | Manual Adjudication |  |  |  |
| --- | --- | --- | --- | --- | --- |
| Natural Language Processing |  | Positive | Negative |  |  |
|  | Positive | 209 (True Positive) | 1 (False Positive) | 210 | **PPV:** 99.5%<br>(95% CI 97.4-99.9%) |
|  | Negative | 7 (False Negative) | 183 (True Negative) | 190 | **NPV:** 96.3%<br>(95% CI 92.6-98.5%) |
|  |  | 216 | 184 | 400 |  |
|  |  | **Sensitivity:** 96.8%<br>(95% CI 93.4-98.7%) | **Specificity:** 99.5%<br>(95% CI 97.0-99.9%) |  | **Accuracy:** 96.6% |

**Table 3:** Performance of the Natural Language Processing Module within the Hypertrophic Cardiomyopathy Output

|  |  | Manual Adjudication |  |  |  |
| --- | --- | --- | --- | --- | --- |
| Natural Language Processing |  | Positive | Negative |  |  |
|  | Positive | 27 (True Positive) | 2 (False Positive) | 29 | **PPV:** 93.1%<br>(95% CI 77.2-99.2%) |
|  | Negative | 2 (False Negative) | 369 (True Negative) | 371 | **NPV:** 99.5%<br>(95% CI 98.1-99.9%) |
|  |  | 29 | 371 | 400 |  |
|  |  | **Sensitivity:** 93.1%<br>(95% CI 77.2-99.2%) | **Specificity:** 99.5%<br>(95% CI 98.1-99.9%) |  | **Accuracy:** 99.0% |

## 4) Discussion

As the information generated through routine clinical encounters continues to grow, there is an important need to ensure that such data are accurately captured and catalogued for research and clinical use. Carefully designed and validated NLP techniques can be deployed on large amounts of EHR text to generate accurate structured output ready for analytic work.^8,11,17-19^ Additionally, open-source NLP – like the module described in this manuscript – has the ability to democratize medical data, enabling researchers and clinicians within a given healthcare system to efficiently analyze free-text clinical data for concepts and diagnoses of interest.

There are a number of novel aspects of our work. First, this NLP was rigorously validated with physician-led adjudication and was determined to be highly accurate. Second, the NLP was developed and validated on echocardiogram reports within a large and diverse healthcare system representing both academic and community-based echocardiography laboratories with a broad range of readers, thereby improving the generalizability and applicability of this work. Finally, given the accuracy of the NLP for categorizing LVH and HCM, the module can be deployed on large collections of echocardiogram reports both for research and broad population-based clinical interventions.

Our work developing this NLP module focused on cardiac structural abnormalities adds to that of other groups focused on developing NLP techniques for cardiovascular and general radiology imaging.^17-20^ Ultimately, the accuracy and portability of any NLP module is subject to the training sets on which it was derived and the programming choices of its designers. This particular NLP was built to accurately phenotype and categorize LVH and HCM for a clinical registry and prospective clinical trials within a large, integrated healthcare system. Unlike methods designed to analyze primary images, our methods provide a more computationally efficient and transparent option for healthcare systems to extract information from echocardiogram reports. Furthermore, a significant benefit of the Canary NLP system^14^ is that researchers and clinician-investigators from other groups can easily modify aspects of its design through its intuitive graphical user interface, **Figure 4**. Unlike other NLP platforms which require a strong background in computer programming (e.g., python), this NLP module can be modified (if desired) and deployed without a significant investment or background in NLP techniques to advance the clinical and research understanding of LVH-associated conditions.

**Figure 4:**
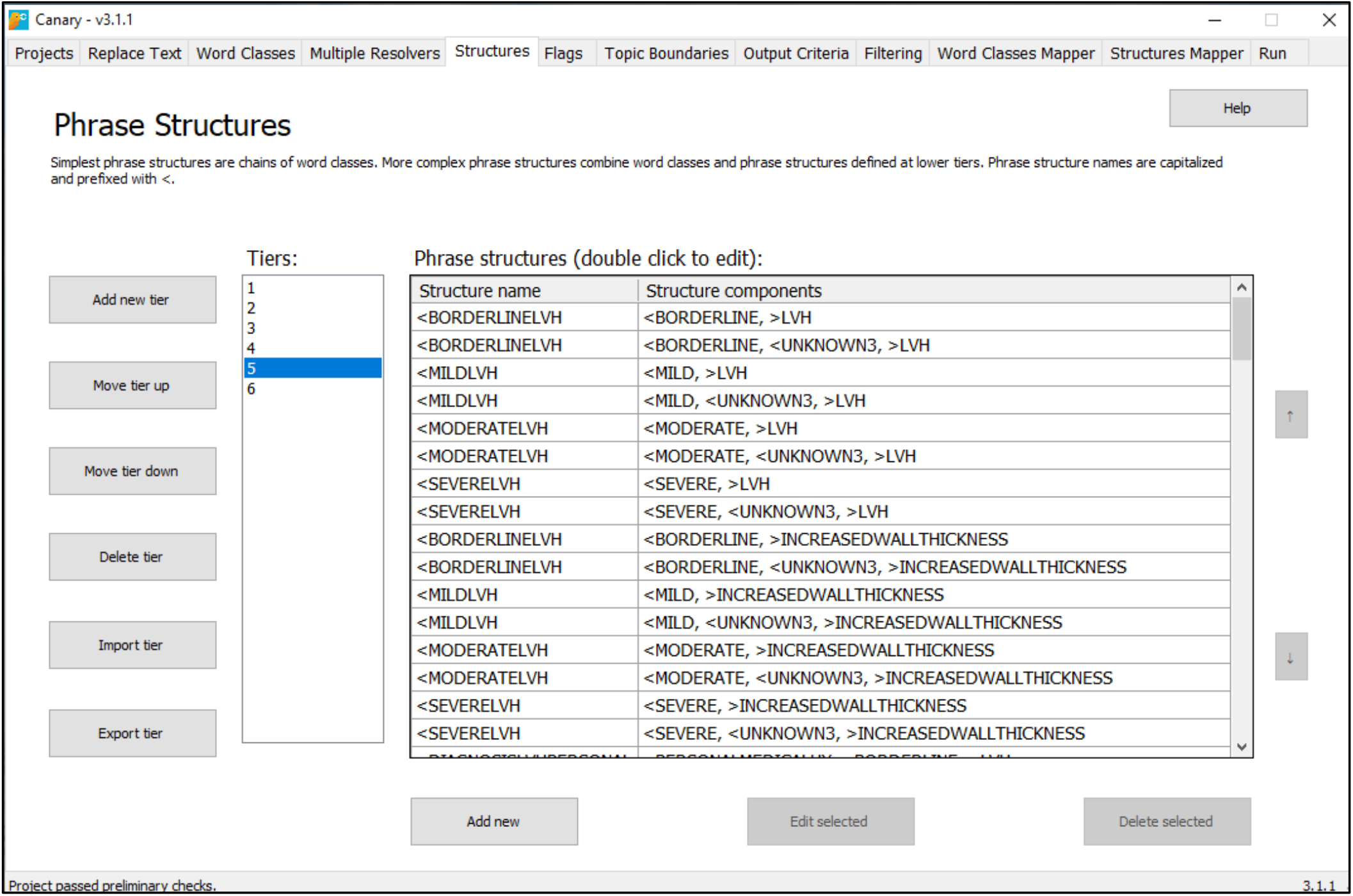
Canary NLP graphical user interface

Despite the accuracy of our validated module, there are some limitations of this work. First, because these modules only extract data based on linguistic descriptors within the echocardiogram reports, it cannot determine if the reported concept is accurate (i.e., if there truly is LVH or findings consistent with HCM). We purposely developed NLP for report texts to facilitate research in which only reports (and not primary images) are available. In the future, if large multicenter repositories of actual echocardiography images become common, other types of artificial intelligence or machine learning methods could be developed and applied directly to images themselves. Finally, because we analyzed echocardiogram reports within the MGB healthcare system from 2017 through 2022, it is possible that the NLP may perform differently on reports from other healthcare systems or on reports prior to 2017. Nevertheless, given the general uniformity of linguistic descriptors of imaging concepts such as LVH and HCM, we would expect similar performance on other sets of echocardiogram reports.

The field of medical informatics is quickly evolving. While machine learning and artificial intelligence techniques are rapidly being developed, they often yield “black box” models without transparency for the results provided and require massive training datasets. Accordingly, we pursued the approach of a human-designed NLP technology that allows the tracing of each step of text analysis and modification of the algorithms to correct errors or add new functionality. With its open-source, user-friendly, and highly accurate phenotyping, we believe that this NLP module will provide the cardiovascular community with an important tool to advance the research surrounding LVH-associated diseases.

## Data Availability

All data produced in the present study are available upon reasonable request to the authors.

https://canary.bwh.harvard.edu/library/

